# The International Concussion and Head Injury Research Foundation Brain health in Retired athletes Study of Ageing and Impact-Related Neurodegenerative Disease (ICHIRF-BRAIN Study)

**DOI:** 10.1101/2022.05.25.22275489

**Authors:** Michael Turner, Cliff Beirne, Antonio Belli, Kaj Blennow, Henrik Zetterberg, Bonnie Kate Dewar, Valentina di Pietro, Conor Gissane, Amanda Heslegrave, Etienne Laverse, Victoria McEneaney, Adrian McGoldrick, James Murray, Patrick O’Halloran, Ben Pearson, Yannis Pitsiladis, Marco Toffoli, Huw Williams, Paul McCrory

## Abstract

**Introduction and aims:** Traumatic brain injury (TBI) is a leading cause of death and disability worldwide. Large registry studies have demonstrated a dose–response relationship between TBI and neurodegenerative disease ; however, disentangling the direct effects of TBI from ageing and/or a progressive neurodegenerative process is problematic. This study is a prospective long-term cohort study to examine a population of retired elite athletes at high risk of concussion and mTBI during their sporting careers compared to age- and sex-matched controls with no history of TBI. The aim is to determine the incidence and risk factors for neurodegenerative disease and/or age-related effects on brain health in this population.

**Methods and analysis:** A population of retired male and female elite athletes and controls aged 40-85 years, will be assessed at baseline and serial time points over 10 years during life using a multi-dimensional assessment including: Questionnaire; SCAT3/5; Neurological and physical examination; Instrumented balance assessment; Computerised neurocognitive screen; Neuropsychological assessment; Advanced MR brain neuroimaging; Visual saccades; Blood workup; Fluid biomarkers; Gut metabolomics; Salivary MicroRNA analysis; Genetic analysis; and where available Brain banking and neuropathology

**Ethics and dissemination:** Ethics approval was granted by St Mary’s University SMEC as well as at the various satellite trial sites. The trial is registered with ISRCTN (BioMed Central) with ID number: 11312093. In addition to the usual dissemination process, this phenotypically well-characterised dataset will reside in a publicly accessible infrastructure of integrated databases, imaging repositories, and biosample repositories and de-identified data will be made available to collaborating researchers.

## INTRODUCTION

### BACKGROUND & RATIONALE

Traumatic brain injury (TBI) is a leading cause of death and disability worldwide. (1) A recent review has highlighted population-based epidemiological evidence linking head injury with dementia. (2) Large registry studies have demonstrated a dose– response relationship between TBI and neurodegenerative disease with a doubling of the risk of dementia following severe injuries, but also a 1.6x increase after mild TBI (mTBI). (3-6) Disentangling the direct effects of TBI from ageing and/or a progressive neurodegenerative process is problematic.

When examining the long-term effects of repeated concussion or repetitive head impacts, there is evidence that some former athletes in contact, collision, and combat sports suffer from depression, cognitive deficits, or mild cognitive impairment later in life. (7-9) Neuroimaging studies show evidence of macrostructural, microstructural, functional, and neurochemical changes in some athletes. (10-19)

Some studies have found an association between these deficits and a history of multiple concussions (7) whereas other studies have not found any relationship. (20) Former high school American football players do not appear to be at increased risk for later life neurodegenerative diseases according to two studies (21, 22) however, an increased risk for neurodegenerative diseases in retired American football (23) and UK professional soccer players (24) is suggested in studies examining death certificates. It is important to appreciate, however, that survey studies of former collegiate (25-27) and professional (28) athletes indicate that the majority of people rate their functioning as normal and consistent with the general population. To date, a cause-and-effect relationship between CTE and concussions or age of exposure to contact sports has not been established. (29-32)

The extent to which repetitive neurotrauma causes static or progressive changes in brain microstructure and physiology and contributes to later life mental health and cognitive problems, is poorly understood, and requires further study.

### TRIAL DESIGN

This study is a prospective long-term cohort study to examine a population of retired elite athletes at high risk of concussion and mTBI during their sporting careers compared to age- and sex-matched controls with no history of TBI.

### TRIAL AIMS AND OBJECTIVES

1. To determine the incidence of and risk factors for neurodegenerative disease in this population
2. To determine the incidence of age-related cognitive change in this population
3. To develop early predictors for neurodegenerative disease in this population

## METHODS

### STUDY SETTING

Volunteers will be recruited by the International Concussion and Head Injury Research Foundation (ICHIRF).

- Phase 1: After online registration of interest at http://www.ichirf.org participants will complete a detailed online questionnaire regarding concussion history, mood, sleep and physical and mental health status. From the database of completed questionnaires, participants will be allocated to groups on the basis of their history of sports related concussion, age and gender. Participants in the concussion group will be matched to control subjects of the same age and gender. A balanced number of female participants to male participants in both concussion/control and age groups will be included in the study.
- Phase 2: Selected participants and controls will undergo an identical screening protocol at the ICHIRF offices in London, UK or at one of the satellite testing centres in Manchester and Dublin, Ireland. The initial cohort screening will commence with those aged > 50 years followed by younger aged groups. The details of the screening program are outlined in the methods section below. Following the completion of all screening elements, the results of the assessment and any findings and/or recommendations will be discussed with the participant and copies of the test results will be made available to them and their general practitioner.
- Phase 3: All participants (including controls) will be serially reassessed at suitable intervals (provisionally every 5 years) using the same multidimensional assessment platform at the ICHIRF offices in London, UK or at one of the satellite testing centres in Manchester and Dublin, Ireland.

### ELIGIBILTY CRITERIA

<u>Inclusion criteria:</u> Participants will be eligible to participate if they

i. have completed the online screening assessment.
ii. have participated in elite sport.
iii. can understand and participate in the testing procedures; and
iv. are able to provide informed consent for participation.

<u>Exclusion criteria:</u> Participants will be ineligible if:

i. For the questionnaire study if they are aged < 18 years
ii. Have a history of previous severe traumatic brain injury,
iii. Are on current psychotropic medication, or
iv. They have a pre-existing medically-diagnosed neurological disorder (e.g., Alzheimer’s dementia, Parkinson’s disease, Multiple Sclerosis, Motor Neuron Disease).
v. The participant is currently enrolled in a disease modifying therapeutic (drug or interventional) trial
vi. Presence of any of the following clinical conditions: Substance abuse within the past year; Unstable cardiac, pulmonary, renal, hepatic, endocrine, hematologic, or active malignancy or infectious disease; AIDS or AIDS-related complex; Unstable psychiatric illness defined as psychosis (hallucinations or delusions) or untreated major depression within 90 days of the screening visit

### FUNDING

The ICHIRF-BRAIN project is funded through a combination of

- <u>Competitive grant funding</u> - EU Erasmus in collaboration with the Galway-Mayo Institute of Technology SCAT Project
- <u>Commercial partnerships</u> - MARKER AG (MicroRNA study)
- <u>Philanthropic support</u> – including Godolphin Racing; the Injured Jockeys Fund; British Association of Sport and Exercise Medicine; the Irish Injured Jockeys; the Professional Footballers Association; the National Football League (US); the Racing Foundation; and private donors.
- <u>Charitable fund raising</u> – Individual contributions from volunteers taking part in the national or local fund-raising initiatives (e.g., The Virgin London Marathon, Vitality 10,000 London Run, Prudential Ride London-Surrey).

### OUTCOMES

#### Primary Outcome

Neurodegenerative disease (e.g., Alzheimer’s Disease) will be a clinical diagnosis supported by neuropsychological, radiological, pathological and/or fluid biomarker changes and in line with international diagnostic criteria. (33-41)

Neurodegenerative diseases of specific interest include Alzheimer’s Disease, Amyotrophic Lateral Sclerosis, Frontotemporal Dementia, Parkinson’s Disease, Lewy Body Dementia, Stroke and Chronic Traumatic Encephalopathy.

#### Secondary Outcomes

Age-related cognitive changes in this population will be a clinical diagnosis supported by neuropsychological, radiological, pathological and/or fluid biomarker changes and in line with international diagnostic criteria. (41-43) Predictors will be based on the study measures listed below.

### SAMPLE SIZE

The aim is to recruit a combined number of 250 participants (concussed and controls), with full phenotypic and investigational workup over a 5-year period, from within the UK, Ireland, and Australia

Incidence of neurodegenerative disease following sport related TBI unknown but in NFL football has been estimated up to 4% of US Professional football participants (McKee et al 2013). The main recruitment group in this study is retired professional jockeys which have a 100x greater risk of concussion/head injury that NFL players based on published study data (Turner et al 2001). Thus 125 retired athlete’s vs 125 controls with a 50/50 male female mix should have sufficient power (0.8) to detect a 0.05 alpha difference between groups

### RECRUITMENT AND ALLOCATION

Volunteers will be recruited by the International Concussion and Head Injury Research Foundation (ICHIRF). After online registration of interest at http://www.ichirf.org participants will complete a detailed online questionnaire regarding concussion history, mood, sleep, and physical and mental health status.

From the database of completed questionnaires, participants will be allocated to groups on the basis of their history of sports related concussion, age, and gender.

### BLINDING

Outcome assessors (imaging, fluid biomarkers, genetics, microRNA, gut metabolomics, visual saccades) will be blinded as to the allocation groups for their assessment and analysis

Emergency unblinding – where abnormal results are noted e.g., on blood tests. The PI will unblind the allocation and notify the participants usual GP re follow up of the abnormal test result. No treatment will be provided.

### PARTICIPANT TIMELINE

Insert Figure 1 about here

**Figure 1:**
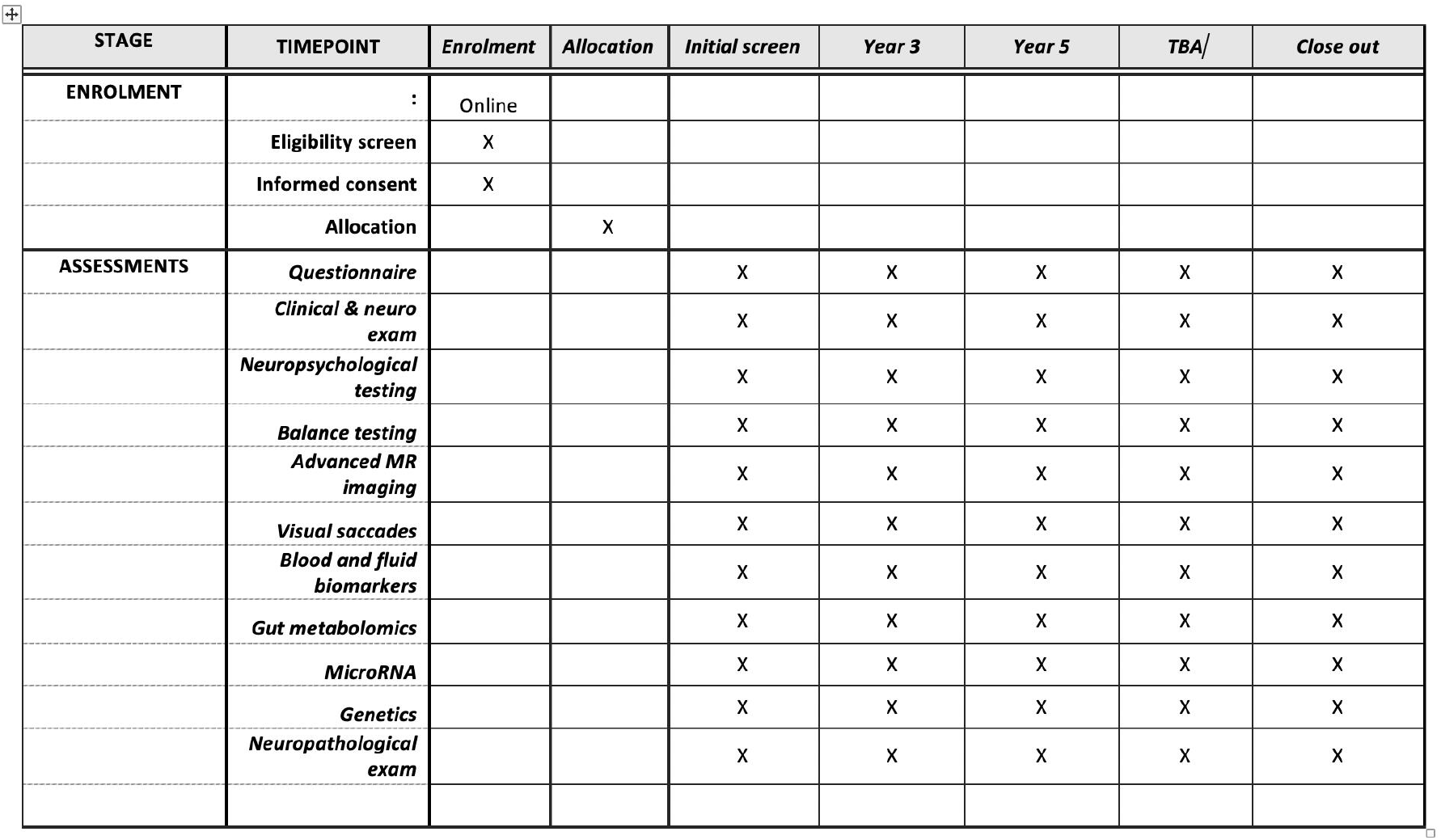
ICHIRF-BRAIN STUDY.

### DATA COLLECTION

During life, a baseline and serial multidimensional assessment on subjects and controls will be done including:

- Questionnaire
- SCAT3/5
- Neurological and physical examination
- Instrumented balance assessment
- Computerised neurocognitive screen
- Neuropsychological assessment
- Advanced MR brain neuroimaging
- Visual saccades
- Blood workup
- Fluid biomarkers
- Salivary MicroRNA analysis
- Genetic analysis
- Brain banking and neuropathology

At baseline and at each assessment visit, participants will complete the following outcome measures:

- ***Questionnaire -*** Participants will complete a questionnaire (appendix 1) which includes questions on past medical history, injury history, concussion history, playing history, sleep, mood. Additional history and information will be sought from the participant’s partner or spouse. Patient reported outcome measures (PROMs) will include DQOL, PDQ, PIMS and MCQ (https://safetyandquality.govcms.gov.au/condition-specific-proms)
- ***Verification –*** On arrival at the screening centre, volunteers will undergo a face-to-face discussion to review the answers provided in the questionnaire (e.g., the number of concussions reported). This data will also be verified by the Consultant Neurologist during screening (see below).
- ***Spouse/partner questionnaire -*** In addition, a partner/spouse questionnaire will be provided on arrival at the screening centre and the answers will be recorded in a face-to face interview with a member of the ICHIRF Staff,
- ***Sports Concussion Assessment Tool 3/5 (SCAT3/5)*** Participants will complete the SCAT3 or SCAT5. (44, 45) The SCAT3/5 includes the following assessments: Glasgow Coma Scale (46), Maddock’s questions (47, 48), Standardized Assessment of Concussion (49, 50), a modified version of the Balance Error Scoring System (mBESS consisting of 3 stances performed on a hard surface) (51), cervical spine examination and modified neurological examination. There are published normative data on this test. (52-55)
- ***Physical and Neurological examination -*** The neurological examination will be performed by a consultant neurologist following a standardized exam protocol. (Appendix 2) Physical examination will include visual acuity and colour vision using Isihara plates, urinalysis and University of Pennsylvania - Brief Smell Identification Test.
- ***Balance Assessment -***In addition to the clinical mBESS assessment (see above), each subject will perform a balance test using the SWAY iPhone app. (56) Sway is a FDA-approved balance test that uses the inbuilt accelerometers in a smart phone or iPad device and objectively measures balance and reaction time
- ***Computerized Neurocognitive Screen -*** Participants will complete the CogState Brief Battery, a validated computerized cognitive assessment (57), which includes four separate tasks: Processing Speed (simple reaction time), Attention (choice reaction time), Learning (visual recognition memory) and One Back (Working Memory test). Participants will perform the test in a quiet room under the supervision of a study investigator. As per test protocol, participants will complete a practice trial for each task before completing the scored test. The primary outcome measure is the speed and accuracy of responses relative to normative data for that age-group.
- ***Neuropsychological assessment -*** Neuropsychological tests for the current study will be selected on the basis of a study into neuropsychological function following repeat concussion in active jockeys. (58) The total administration time for the neuropsychological battery will be approximately 60 minutes and will be performed by a consultant clinical neuropsychologist. All neuropsychological tests will be administered and scored according to standardised instructions. The following neuropsychological domains assessed: premorbid function (Test of Premorbid Function). (59); vocabulary and verbal ability (Vocabulary subtest from Wechsler Adult Intelligence Scale-Fourth UK Edition WAIS-IV) (60); auditory verbal short term and working memory (Digit Span subtest from the WAIS-IV) (60); processing speed (Symbol Digit Modalities Test) (61); and the Speed of Comprehension Test) (62); verbal learning and memory (California Verbal Learning Test II) (63); response inhibition (Stroop) (64), visual scanning and response alternation (Colour Trails Test) (65); and fluency across both semantic and letter conditions. Administration of the digit span subtest from the WAIS-IV will allow the use of an embedded measure (Reliable Digit Span) sensitive to the application of cognitive effort. Preliminary analysis will compare performance on neuropsychological composites, the number of abnormal scores and performance on individual tests between concussion and control groups. Future analyses will investigate the relationship between neuropsychological test performance and co-morbid factors such as mood, a history of learning difficulties and/or attention deficit hyperactivity disorder, substance use, and pain; and between neuropsychological test performance and sports related factors such as number of concussions, age at first concussion, and type of sport. Finally, relationships between neuropsychological test performance and imaging, neurology, balance and eye movements will also be explored
- ***Advanced MR brain imaging -*** Anatomical and functional magnetic resonance imaging (MRI) will be acquired for all subjects. MRI studies will be performed using a 3.0 Tesla Siemens scanner using a 32-channel head coil and will require a minimum of 45 minutes to complete. The image sequences will include:
- Structural scans, consisting of 3D T1-weighted (T1w) high-resolution sequences and fluid-attenuated inversion recovery (FLAIR) sequences with whole brain coverage. The T1w and FLAIR sequences will permit detection of any underlying structural lesion. Volumetric analysis will be performed using the 3D T1-weighted high-resolution sequences Magnetisation Prepared Rapid Gradient Echo (MPRAGE) sequence. Data measurement and analysis will be as per Guo et al. (1).

Analysis of T1w MRI shall be undertaken to detect subtle changes in brain morphology and morphometry related to mild TBI, such as by voxel-based morphometry for whole-brain analysis, volumetric changes with Geodesic Information Flows and FSL-FIRST for analysis of the subcortex. We shall also ascertain the predicted brain age of all participants using BrainAgeR. Utilising both the T1w and FLAIR sequences shall also permit detection of white matter hyperintensities across the cohort.

- Diffusion weighted imaging (DWI) (B3000), which will allow for mapping of white matter tracts in the brain. Analysis will be undertaken for both tract based spatial statistics and tractography, such as with constrained spherical deconvolution (CSD), a method robust to crossing fibres.
- T2-relaxometry and susceptibility weighted imaging sequences, which allows detection of regional grey and white matter changes that may reflect long-term changes in the brain following mild TBI, including the presence of haemosiderin staining and microhaemorrhage.
- Resting-state functional MRI (rs-fMRI), acquired by blood oxygenation level dependent (BOLD), will be acquired to assess both resting state functional activity and functional connectivity, and its relationship to mild TBI. This shall include investigating for brain connectivity network differences between the cohort, its relationship to other collected patient parameters including biomarkers. Graph theory metrics of this shall also be collected, including node centrality measures.

Analysis of the scans will be performed using standard and well-validated statistical techniques, including by non-parametric randomised permutation testing with appropriate statistical correction for multiple comparisons, and Bayesian regression models.

- ***Visual Saccadic testing -*** Saccadic latencies (reaction times) will be recorded using a portable, microminiaturised head-mounted saccadometer, in accordance with a standard published methodology. (49) Three lasers projected high-contrast red targets in a horizontal line at -10°, 0° and +10° on a wall in front of the seated participant. Each trial begins with the central target illuminated. After a random delay of 0.5–1.5s, this jumps 10° to the left or right randomly. Participants are instructed to follow the target with their gaze and 200 saccades will be recorded, taking around 7 minutes. The device records saccadic latency using scleral infrared oculometry, with automatic deletion of blinks, movements in the wrong direction and those with an abnormal velocity profile. Each participant’s saccadic latency distribution will be analysed using custom-built software, which calculates best-fit parameters using an established model of saccadic latency, as previously described. (67, 68)
- ***Blood testing -*** Participants in this study will have blood drawn and labeled at baseline and each assessment time point. Samples will be anonymised prior to analysis. Professional phlebotomists will draw all blood. Routine blood workup will be analysed under the supervision of The Doctors Laboratory Ltd, 60 Whitfield Street, London, W1T 4EU, UK. The blood screen includes: Full blood count + 5-part differential; erythrocyte sedimentation rate; c-reactive protein; sodium; potassium; chloride; bicarbonate; urea; creatinine; bilirubin; alkaline phosphatase; aspartate transaminase; alkaline transaminase; creatine kinase; lactate dehydrogenase; gamma glutamyl transferase; total protein; albumin; globulin; calcium; phosphate; uric acid; random blood glucose; cholesterol; high density lipoprotein; low density lipoprotein; triglycerides; serum iron; total iron binding capacity; ferritin; vitamin D; blood group; free thyroxine (T4); thyroid stimulating hormone; growth hormone; cortisol; prolactin; coeliac disease profile (tissue transglutaminase (IgA), HLA DQ2/DQ8, total immunoglobulin A). In females only, luteinizing hormone and follicle stimulating hormone. In males only, prostate profile -total prostate specific antibody, free prostate specific antibody, calculated ratio.
- ***Fluid Biomarkers studies -*** Participants in this study will have 5ml Serum and 5ml plasma drawn, labelled, stored in cryovials of 0.5ml and frozen at -80**°**C in liquid nitrogen at baseline and at each assessment time point. Samples will be anonymised prior to analysis. Professional phlebotomists will draw all blood. Future consideration to performing fluid biomarkers on CSF will be undertaken. Samples will be batch-tested using ultrasensitive single molecule array (Simoa) methods (Quanterix, Billerica, MA) (69, 70) for the following biomarkers: neurofilament light polypeptide (NFL), tau, ubiquitin carboxyl-terminal hydrolase isoenzyme L1 (UCH-L1), glial fibrillary acidic protein (GFAP), Aβ40 and Aβ42. Genotyping the apolipoprotein E (*APOE*) ε4 allele will also be performed. Neuron-specific enolase (NSE) and S100B will be measured using immunoassays with electrochemiluminescence detection.
- ***MicroRNA Study*** – Saliva samples will be obtained from participants and subject to Next Generation Sequencing (NGS) analysis for the identified miRNAs. NGS sequencing libraries will be prepared, quantified and sequenced for all samples. The collected reads will be subjected to quality control, unique molecular index-based correction (to remove PCR replicates), alignment and downstream analysis. Identified miRNA’s will be validated by qPCR, in situ hybridization or miRNA inhibition.
- ***Genetics* -** The advantages of using saliva for whole genome sequencing (WGS) include -the ease in obtaining samples non-invasively from participants, the convenience in mailing saliva collection kits and the long-term stability of saliva samples at room temperature. However, as saliva samples have substantially lower DNA yield than blood, and are prone to microbial contamination, a carefully standardised saliva collection protocol is essential for saliva DNA to meet the stringent QC metrics needed to generated good quality WGS data. Saliva samples will therefore be collected from participants using the Oragene DNA Self-Collection kit (tube format OG-500; DNA Genotek Inc., Kanata, Ontario, Canada) and used for DNA extraction. Each sample will be bar-coded, temporarily stored at room temperature and subsequently transferred to a central laboratory for DNA extraction, biobanking and subsequent analysis (71). DNA will be extracted from a 500 μl aliquot from the Oragene DNA/saliva Self-Collection kits in accordance with the manufacturer’s instructions. Extracted samples will be stored at −20°C prior to NGS. WGS will be performed using DNBSEQ-G400RS (BGI, Shenzhen, China) to a target average coverage depth of 30x and a read length of 150 bp. WGS will be used to determine and compare common and rare single nucleotide variants (SNV) and copy number variants (CNV) between cases and controls. (71)
- ***Neuropathological brain examination -*** Participants and control subjects will be given the opportunity to enrol in the brain bank program, which requires specific informed consent through University College London. If willing to participate, the names and contact details will be forwarded to the Queen Square Brain Bank (QSBB) for neurological disorders coordinator who will make contact and provide additional information as required. All further dialogue with the volunteer will be coordinated by the QSBB who will enrol the participant in the brain bank program. In the event of the participant dying, the QSBB coordinator will make the necessary logistical arrangements with the family, hospital, or funeral director for harvesting brain tissue for detailed neuropathological analysis and tissue preservation for future research. All material stored at the QSBB is under HTA licence and any tissue used for research will have ethical approval obtained from the National Research Ethics Service Committee London. The neuropathological assessment, including staining methods, anatomical sampling sites and diagnostic criteria will be in accordance with a recent NIH consensus conference.

### DATA MANAGEMENT

Each individual will be given a unique data identification number of Individuals will not be identifiable from the data.

Data will be double entered, and range checked into the relevant database spreadsheet and further checked by the PI and study statistician for completeness

### STATISTICAL METHODS

The main aim is to examine the characteristics of concussed and control subjects. Initially, descriptive data for each variable was calculated. The descriptors will vary depending upon the data type. For continuous variables mean (and SD) will be computed. If the variables are categorical, medians will be calculated for ordinal variables, and proportions for nominal variables. Significance will be set a priori at P<0.05. All estimates will be accompanied by a 95% confidence interval. All computations will be conducted using R statistics for computing. In addition to the base package, the MASS, Robustbase, EpiR, Psych, Effsize, lsr, desctools, and corrplot packages will be employed.

- **Primary Outcome Analysis**: As the study is observational, volunteers being either a case of concussion or a control, regressions will be used for analysis. The outcome variable will determine the specific regression; binary variables, logistic regression; continuous variables, linear regression; ordered logistic regression ordinal variables. For some variables robust regressions will be needed to account for and reduce the weight (influence) of outlying observations. <u>Robust regression will use MM estimators to decrease inefficiency that leads to loss of power.</u> (72) Regardless of the type of regression, Age and gender will be included in the analyses, as the two potential confounders.(73)
- **Additional Analyses:** To address potential underlying factors, the neuropsychological measures and the biomarkers data sets will be examined using exploratory factor analyses (EFA). <u>The technique is designed to reveal any underlying relationships between measured variables.</u> The resulting EFA composite scores from the regression function will be entered into any subsequent regression analysis. For neuropsychological analysis three factors were established: Combined Cognition index (CCI), Cognition Memory Index (C-MI) and Cognition – Executive/Speed Index (C-ESI). For biomarkers factors were identified including Tau, Abeta40, Abeta42, NFL, GFAP and UCH-L1.
- **Treatment of Missing Data**: The treatment of missing data values depended upon the type of data. For continuous variables, volunteers were assigned the mean value score for that variable. For categorical variables, the median was applied.

### DATA MONITORING

No formal Data Monitoring Committee is required as this is an observational study

### INTERIM ANALYSIS

An interim analysis of the result is planned at the conclusion of Phase 1 (screening of initial cohort) to determine the usefulness of the outcome measures and whether additional outcome measures need to be considered. As this is not an interventional study, the study will not be ‘terminated’ on the basis of an interim analysis

### SAFTETY/HARMS

There are no anticipated major adverse risks with the testing. If the participants become upset about the questioning or the procedures the testing will stop. Professional phlebotomists will draw all blood samples. The questionnaires being used have been used in research previously. They are valid and dependable, and it is not anticipated that adverse reactions will take place.

Each volunteer will get an individual report within 30 days outlining the results of their screening and making recommendations about any further action that is required.

### AUDITING

The PI will audit the various components of the trial conduct and will conduct regular site visits to all test sites and analysis laboratories to check compliance with study protocols

### RESEARCH ETHICS APPROVAL

All participants will provide written informed consent as well as written informed consent from their next of kin where appropriate.

Ethical approval for the study was obtained as follows (including protocol amendments):

<u>St Mary’s University SMEC</u>

- Study outline and questionnaire – 01/06/2015 (no reference number)
- Screening protocol – 27/10/2015 Reference: SMEC-2015-16-53
- Brain donation – 12/06/2017 Reference: SMEC-2016-17-115

<u>University of Birmingham</u>

- Saliva study – 22/09/2017 Reference: 17/EE/0275 IRAS project ID – 216703

<u>Beacon Hospital, Dublin, Ireland</u>

- Dublin satellite test site – 10/10/2019 Reference: BEA0130

### CLINICAL TRIAL REGISTRY

The trial is registered with ISRCTN (BioMed Central) with ID number: 11312093

### INFORMED CONSENT PROCESS

All participants will provide written informed consent as well as written informed consent from their next of kin where appropriate as per the Ethic Committee approvals.

### CONFIDENTIALITY

Each individual will be given a unique data identification number of Individuals will not be identifiable from the data. The data will only be accessed by principal investigators and data will be stored in hard copy and electronically at the offices of ICHIRF in UCL London, at the Beacon Hospital, Dublin and backed up at the University of Melbourne.

### OPEN ACCESS TO DATA

One of the fundamental aims of the ICHIRF project is to enable data sharing, prevent duplication of efforts and foster collaboration among research teams in order to accelerate research in TBI. This phenotypically well-characterised dataset will reside in a publicly accessible infrastructure of integrated databases, imaging repositories, and biosample repositories. We believe that such datasets only serve their intended purposes with a robust, transparent, and open-access data sharing plan.

Approximately 12 months after the end of the initial phase of the project, de-identified data will be made available to collaborating researchers involved in the project. Twelve months after publication of the results, data will be open to all qualified and approved researchers. We will develop policies and procedures to allow us to collaborate and share data throughout the study to advance knowledge in TBI.

### ANCILLARY AND POST TRIAL CARE

As this is an observational study, no harms are expected for the trial subjects. In all cases, a copy of the results is given to the subject and, with their permission, their usual GP

### DISSEMINATION POLICY

#### Participants

All volunteers will receive a detailed report, including the results of any tests conducted, within 30 days of the screening. This will include recommendations regarding any further action that may be required as a result of any abnormal findings (e.g., in the event of a raised cholesterol, the volunteer would be advised to consult his/her GP and recommended to have the test repeated).

#### Publication

De-identified trial data will be presented at conferences, seminars and published in the medical literature.

#### Open Access

At the end of the initial phase of the project, de-identified data will be made available to collaborating researchers involved in the project. After publication of the results, data will be open to all qualified and approved researchers.

### AUTHORSHIP

Authorship will be open to all contributors involved in the study who fulfil the ICMJE 2018 authorship requirements namely

- Substantial contributions to the conception or design of the work; or the acquisition, analysis, or interpretation of data for the work; AND
- Drafting the work or revising it critically for important intellectual content; AND
- Final approval of the version to be published; AND
- Agreement to be accountable for all aspects of the work in ensuring that questions related to the accuracy or integrity of any part of the work are appropriately investigated and resolved.

### REPRODUCIBILITY OF RESULTS

As discussed above, following the conclusion of the trial and publication of the results, data will be open to all qualified and approved researchers.

### EXPECTED OUTCOMES

1. Provide detailed information on the prevalence of mental health issues, cognitive impairment, and/or neurodegenerative disease in a cohort of retired professional athletes vs. control subjects
2. Identify risk factors in this cohort that may lead to or predict the severity of mental health issues, cognitive impairment, and/or neurodegenerative disease in this population
3. Identify the health and personal impact of TBI-related sequelae
4. Establish the role of advanced multimodal assessments in this setting.
5. Improve TBI-related sequelae taxonomy for future targeted clinical treatment trials
6. Identify new diagnostic and prognostic markers and refine outcome assessments for this population
7. Create an open access database enabling research collaboration and data sharing capability
8. Disseminate the research data to community athletes

## DISCUSSION

Recent studies have suggested an association between contact sport participation and an increased risk of late neurodegenerative disease but separating the effects of brain trauma from ageing, unrelated mental health or neurodegenerative disease, is problematic. To assess the hypothesis that contact sport participation is linked to an increased incidence of dementia, prospective longitudinal studies such as the ICHIRF-BRAIN study are critical.

## Data Availability

All data is under collection and will be available as outlined in the paper

https://ichirf.org/

## Acknowledgements

The ICHIRF-BRAIN Study would not be possible without the input of the ICHIRF Project Manager, Pippa Theo, and the Queen Square Brain Bank Coordinator, Karen Shaw.

## APPENDIX 1 NEUROLOGICAL EXAMINATION

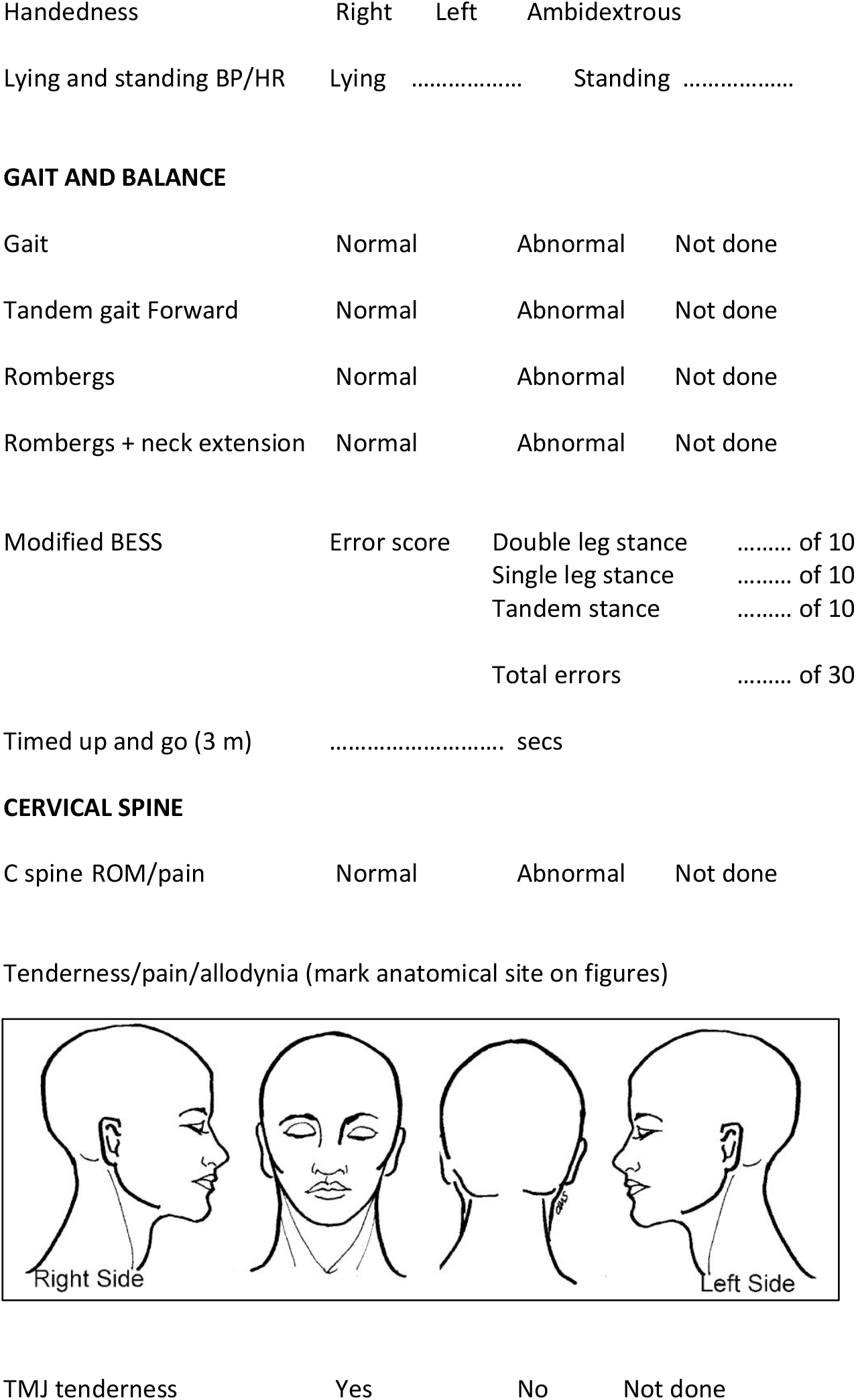
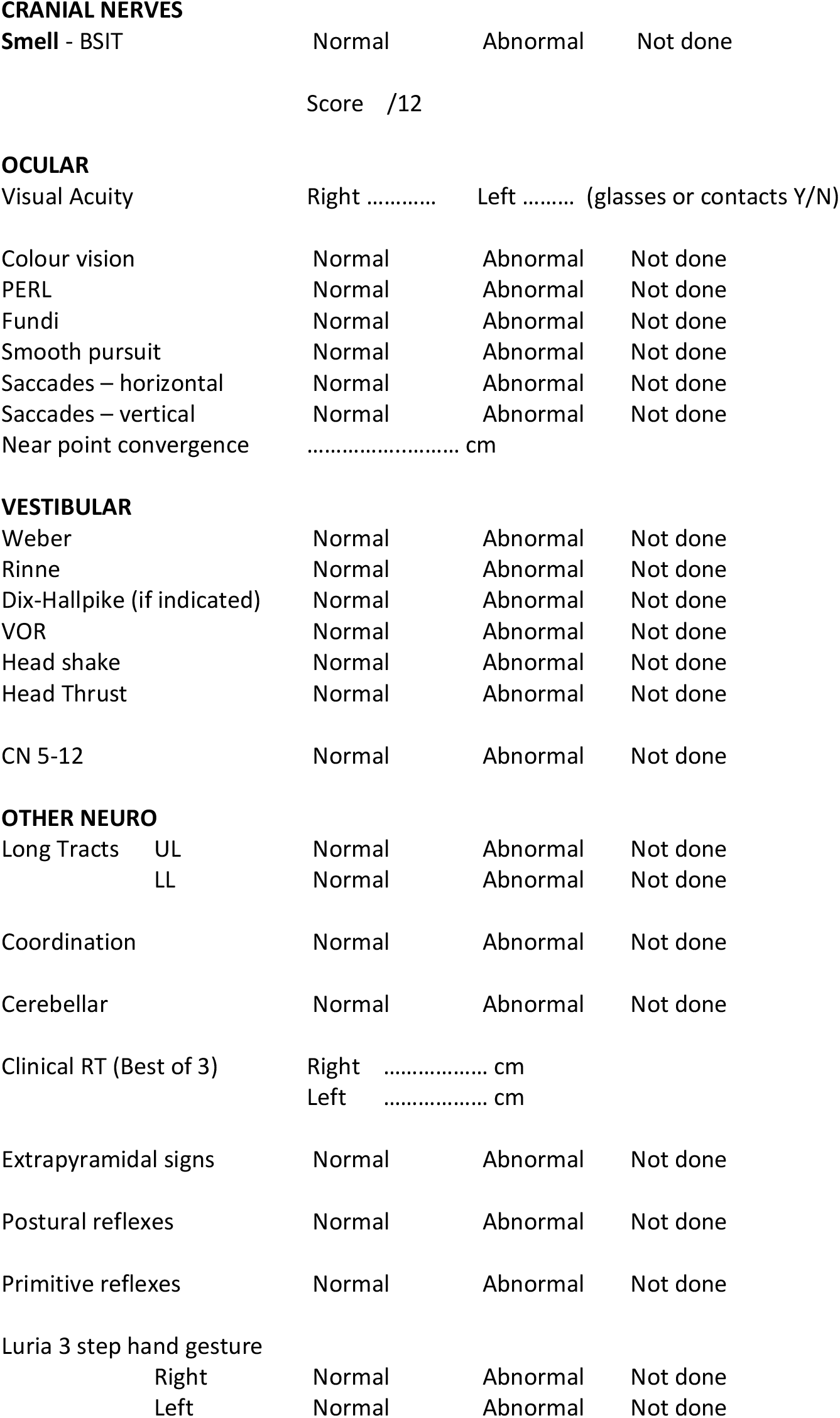

